# Statin Therapy and Stroke Risk in Patients with Hypercholesterolaemia: A population-based longitudinal study using electronic health records in South London

**DOI:** 10.1101/2025.03.10.25323670

**Authors:** Marc Delord, Abdel Douiri

## Abstract

**Background:** We aim to evaluate the impact of statin prescription on stroke risk in patients with hypercholesterolaemia and to assess disparities in statin prescribing.

**Methods:** We analysed electronic health records from patients with hypercholesterolaemia, registered in 41 general practices in south London between 2005 and 2021. The cause-specific hazard ratio of statin prescription on stroke, adjusted for patients’ sociodemographic characteristics and stroke risk factors (smoking ever, hypertension, and diabetes), was estimated using a time-varying exposure Cox proportional hazards model stratified by history of heart diseases. The association between statin prescription and patients’ sociodemographic characteristics was evaluated using a logistic regression.

**Results:** Of the 849,968 registered patients, 166,124 (19.5%) had records of hypercholesterolaemia. Among them, 33.5% were prescribed statins, 2.6% had a record of stroke, and 50.6% were female, 31.7%, 16.2% and 8.9% had records of hypertension, diabetes and history of heart diseases respectively. In a Cox model stratified by history of heart diseases, statin prescription was associated with a reduced hazard of stroke (cause-specific hazard ratio: 0.74; 95% confidence interval (CI): 0.68–0.80, p<0.001), with follow-up administratively censored at 79 years (n=161,527; 97.2%). Statins were less likely prescribed to female patients and patients of Black ethnicity (odds-ratio: 0.70, 95% CI: 0.68-0.72, p<0.001 and odds-ratio: 0.82, 95% CI: 0.79-0.85, p<0.001, respectively).

**Conclusions:** Statin therapy prescription is associated with reduced stroke risk in patients with hypercholesterolaemia, yet it was under-prescribed to women and patients of Black ethnicity, highlighting avoidable disparities in preventive care.

## Background

The relationship between hypercholesterolaemia and stroke has been described as complex [1]. This complexity is due to interacting factors such as cholesterol fractions [2], stroke subtypes [2–4], patients’ age [5], and the confounding effect of lipid-lowering therapies, particularly statins. For instance, the risk of stroke in patients with familial hypercholesterolaemia (FH), which is associated with high low-density lipoprotein cholesterol (LDL-C) [6], is significantly reduced by the use of statins [7], and recent population-based studies did not report an excess risk of stroke in patients with FH [8, 9]. Despite initial concerns regarding the paradoxical association between cholesterol levels and haemorrhagic stroke [3, 10], statins are considered safe and effective to reduce total stroke risk [5, 11–14].

Importantly, statins reduce the risk of major vascular events, including stroke, irrespective of patients’ age, sex, or cardiovascular risk [12, 13], lower risks being associated with more intensive regimens [13]. Statins are also considered remarkably safe drugs, with possible risks of haemorrhagic stroke being outweighed by the protective effect against ischaemic stroke and coronary artery disease [15]. Concerns about statin-related muscle pain were explored in randomised trials, which showed no significant difference in muscle symptoms between statin and placebo groups [16, 17]. Other significant but rare adverse effects of statins include new-onset diabetes [18] or muscle damage such as myopathy or rhabdomyolysis [15].

Overall, lifetime statin regimens have proven to be cost-effective [19] and are currently recommended for patients with a history of cardiovascular diseases, or in primary prevention based on factors such as pre-treatment cardiovascular risk, LDL-C levels, or comorbidity [20–23]. Since 2014, the National Institute for Health and Care Excellence (NICE) guidelines for primary prevention recommended statin prescription in patients aged 40 and older with a 10-year cardiovascular risk above 10% (QRISK2 > 10%) [24]. This threshold was set to prevent cardiovascular events in patients considered at moderate risk. These guidelines also suggested considering statins if other factors such as family history, socioeconomic factors, or patient preference might indicate an underestimated cardiovascular risk. The 2023 update reinforces the importance of shared decision-making, allowing statin use even for patients with a risk lower than 10% if the patient has an informed preference or if the risk might be underestimated. This update also recommends using the QRISK3 tool for cardiovascular risk assessment, which accounts for additional risk factors such as chronic kidney disease, severe mental illness, and migraine, and emphasises flexibility and patient autonomy in making treatment decisions [22, 25].

Despite the favourable benefit–risk profile of statins, concerns exist regarding treatment adherence in primary or secondary cardiovascular disease prevention [26]. Consequently, optimal lowering of LDL-C is not achieved within two years in over half of the patients initiated on statin therapy, significantly elevating their risk of future cardiovascular disease [27]. On the other hand, the effectiveness of statins for primary stroke prevention [28], even in patients without substantial cardiovascular risk factors [13], suggests a potential for broader use of statins in primary prevention of stroke in patients with hypercholesterolaemia.

In line with these observations, our objective in this study is to better understand the association between the onset of stroke and statin prescription in patients with hypercholesterolaemia, using routinely collected electronic health records in a multiethnic, youthful, and urban population. Accordingly, we have estimated the impact of statin prescription on stroke hazard in patients with hypercholesterolaemia. The association between statin prescription and patients’ sociodemographic characteristics was also evaluated.

## Methods

### Study participants

The study was conducted in the Lambeth borough in South London, characterised by a multiethnic, youthful, and deprived population. We utilised primary care electronic health records from adult patients aged over 18 and registered in one of the 41 Lambeth general practices between April 2005 and April 2021. The proportion of patients dropping out from the anonymised data-sharing scheme was 3.2%. These patients were not included in the study.

### Primary care data

Electronic health records included the date of diagnoses of 18 long-term conditions coded in accordance with the Quality and Outcomes Framework (QOF), based on QOF38 definitions [29]. Long-term conditions included stroke, hypertension, diabetes, hypercholesterolaemia (total cholesterol > 5.0 mmol/L), atrial fibrillation, heart failure, myocardial infarction, chronic kidney disease stage 3 to 5, and transient ischaemic attack. Other information included gender (self-ascribed), ethnicity (Asian ethnicity, Black ethnicity, mixed ethnicity, other/unknown ethnicity, and White ethnicity), ‘ever smoking’ status, and the Index of Multiple Deprivation (IMD) 2019 (from 1-most deprived to 5-least deprived, based on seven domains of deprivation including income, employment, education, health, crime, housing, and quality of living environment). From available records, long-term conditions eligible for primary prevention include diabetes and chronic kidney disease stage 3 to 5. Likewise, patients eligible for secondary prevention are patients with a history of cardiovascular disease, defined as coronary heart disease, heart failure, stroke, peripheral arterial or vascular disease, myocardial infarction, atrial fibrillation, or transient ischaemic attack. History of heart disease was defined by at least one record of myocardial infarction, heart failure, coronary heart disease or atrial fibrillation. Likewise, history of cardiovascular diseased was defined by history of heart disease or at least one record of transient ischemic attack, peripheral arterial/vascular disease.

### Statistical analyses

Patients were followed until stroke occurrence (primary outcome), death (competing event), or censoring at last contact for patients who remained stroke-free and alive. Long-term conditions, sociodemographic covariates, and other variables were displayed as frequencies or median and interquartile range as appropriate.

The cause-specific hazard of stroke in patients with hypercholesterolaemia, in relation to statin prescription, sociodemographic covariates, and stroke risk factors, was evaluated using a multivariable Cox proportional hazards model [30–32]. In this model, statin prescription was modelled as a time-dependent covariate, with patients considered unexposed until the date of their first statin prescription. The proportional hazards assumption regarding variables fixed at baseline was assessed using the Schoenfeld residual-based test. Patients’ follow-up was administratively censored to ensure that the proportional hazards assumption was satisfied. The association between statin prescription, sociodemographic covariates, and stroke risk factors in patients with hypercholesterolaemia was evaluated using multivariable logistic regression. This model was also adjusted for age, with follow-up censored at the age of stroke for patients who experienced a stroke and at the age at last follow-up for other patients. Model calibration was visually assessed by plotting predicted probabilities of statin prescription against observed proportions per decile of predicted probabilities and numerically evaluated using the Brier score, which reflect both the calibration and discriminative ability of the model. This score ranges from 0 (perfect accuracy) to 0.25 for a non-informative model when the outcome prevalence is 50% [33]. The area under the receiver operating characteristic curve (AUC), which quantifies discriminative ability of the model was also evaluated [34].

Patients with missing covariates were excluded from models where these covariates were used as confounders.

All computations were carried out using the R language and environment for statistical computing (version 4.3.0 (2023-04-21)) [35].

### Ethics

This study was conducted in accordance with the Declaration of Helsinki. Data were provided by the Lambeth DataNet upon approval for the analysis of fully anonymised data by the Lambeth Clinical Commissioning Group. All patients were informed through ‘Fair Processing Notices’ of the potential use of collected electronic health records in ‘secondary data analysis’ and were given the option to opt out of the data-sharing scheme [36]. Accordingly, this study was exempt from ethics committee approval and individual consent requirements as per the National Institute for Health (NIH) Health Research Authority (HRA) guidelines for research using anonymised primary care data in the United Kingdom.

### Patient and public involvement

This research project actively involved patients and the public at several stages, including study design and dissemination strategies. Feedback was gathered from two Patient and Public Involvement groups, organised by the NIHR Biomedical Research Centres, including stroke and cardiovascular patient groups. These consultations were invaluable in shaping our approach to data use, confidentiality, and research ethics.

To ensure transparency and better inform the public about Lambeth DataNet research outcomes, the results of this research will be disseminated widely. Efforts will involve sharing findings with the public through accessible platforms, including community websites and publications, as recommended by the Lambeth DataNet engagement groups. The results will also be communicated to local community groups, healthcare professionals, and Patient and Public Involvement groups to continue the discussion and identify further areas of research.

## Results

### Patient characteristics

Of 849,968 registered patients, 166,124 (19.54%) had hypercholesterolaemia and 9,847 (1.15%) had a record of stroke. Characteristics of patients with hypercholesterolaemia are summarised in Table 1. Briefly, 8.3%, 21.9%, 3.7%, and 54.4% self-identified as being of either Asian, Black, Mixed, or White ethnicity, and 11.6% were of other or unknown ethnicity. Female gender (self-ascribed) represented 50.6%. Gender was not declared by three patients. The IMD was available for 164,322 (98.9%) patients. Patients of White and Asian ethnicity had similar IMD profiles, whereas patients of Black ethnicity were overrepresented in IMD quintiles 1 and 2 (most deprived) and conversely less represented in quintiles 4 and 5 (least deprived).

**Table 1.** Patient characteristics according to statin prescription.

| Patient characteristics |  | Statins |  | Total |
| --- | --- | --- | --- | --- |
|  |  | Not prescribed | Prescribed |  |
| Total N (%) |  | 110522 (66.5) | 55602 (33.5) | 166124 |
| Age at end of follow-up | Median (IQR) | 50.7 (19.8) | 67.2 (19.5) | 56.2 (22.2) |
| Gender | Male | 53001 (48.0) | 29116 (52.4) | 82117 (49.4) |
|  | Female | 57518 (52.0) | 26486 (47.6) | 84004 (50.6) |
| Ethnicity | White | 61809 (55.9) | 28584 (51.4) | 90393 (54.4) |
|  | Black | 22568 (20.4) | 13807 (24.8) | 36375 (21.9) |
|  | Asian | 8014 (7.3) | 5857 (10.5) | 13871 (8.3) |
|  | Mixed | 4323 (3.9) | 1807 (3.2) | 6130 (3.7) |
|  | Other | 3329 (3.0) | 1334 (2.4) | 4663 (2.8) |
|  | Unknown | 10479 (9.5) | 4213 (7.6) | 14692 (8.8) |
| IMD local quintile | 1 (most deprived) | 22683 (20.8) | 13134 (23.9) | 35817 (21.8) |
|  | 2 | 22449 (20.5) | 12047 (21.9) | 34496 (21.0) |
|  | 3 | 19876 (18.2) | 9253 (16.8) | 29129 (17.7) |
|  | 4 | 20899 (19.1) | 9528 (17.3) | 30427 (18.5) |
|  | 5 (least deprived) | 23346 (21.4) | 11107 (20.2) | 34453 (21.0) |
| Age at record of Hypercholesterolaemia | Median (IQR) | 43.5 (16.4) | 55.3 (18.3) | 47.2 (18.4) |
| Stroke | No | 108813 (98.5) | 52921 (95.2) | 161734 (97.4) |
|  | Yes | 1709 (1.5) | 2681 (4.8) | 4390 (2.6) |
| Hypertension | No | 91292 (82.6) | 22199 (39.9) | 113491 (68.3) |
|  | Yes | 19230 (17.4) | 33403 (60.1) | 52633 (31.7) |
| Diabetes | No | 105454 (95.4) | 33750 (60.7) | 139204 (83.8) |
|  | Yes | 5068 (4.6) | 21852 (39.3) | 26920 (16.2) |
| CKD 3-5<sup>a</sup> | No | 107673 (97.4) | 46924 (84.4) | 154597 (93.1) |
|  | Yes | 2849 (2.6) | 8678 (15.6) | 11527 (6.9) |
| Smoking ever | No | 54301 (49.1) | 22363 (40.2) | 76664 (46.1) |
|  | Yes | 56221 (50.9) | 33239 (59.8) | 89460 (53.9) |
| Obesity (moderate to severe) | No | 78984 (71.5) | 28701 (51.6) | 107685 (64.8) |
|  | Yes | 31538 (28.5) | 26901 (48.4) | 58439 (35.2) |
| History of heart disease<sup>b</sup> | No | 107768 (97.5) | 43623 (78.5) | 151391 (91.1) |
|  | Yes | 2754 (2.5) | 11979 (21.5) | 14733 (8.9) |
| History of cardiovascular disease<sup>c</sup> | No | 107268 (97.1) | 41381 (74.4) | 148649 (89.5) |
|  | Yes | 3254 (2.9) | 14221 (25.6) | 17475 (10.5) |
| Diabetes<sup>*d</sup> or CKD 3-5 | No | 103913 (94.0) | 30042 (54.0) | 133955 (80.6) |
|  | Yes | 6609 (6.0) | 25560 (46.0) | 32169 (19.4) |

<sup>a</sup> Chronic Kidney Disease Stage 3-5<sup>b</sup> Myocardial infarction, Heart Failure, Coronary Heart Disease, Atrial Fibrillation<sup>c</sup> History of Heart Disease, Transient Ischemic Attack, Peripheral Arterial/Vascular Disease<sup>d</sup> Diabetes after 40 years old or for more than 10 years

### Risk factors, stroke, and statin prescription

Among patients with hypercholesterolaemia, 33.5% were prescribed statins versus 1.3% in other patients, and 4,390 patients (2.6%) had a record of stroke. These patients represented also 44.6% of all patients with stroke in the registry. Hypertension (67.5%) and diabetes (29.7%) were the two major metabolic stroke risk factors recorded for these patients, along with a history of heart disease (25.1%) and atrial fibrillation (12.6%).

Patients of Black ethnicity were overrepresented among patients with a record of stroke (26.6% vs. 21.9%), and conversely, patients of White ethnicity were slightly underrepresented (52.4% vs. 54.4%) (Additional file 1: table S1) .

Statins were prescribed in 33.5% of patients with hypercholesterolaemia and 32.9% of patients with stroke. Among patients with hypercholesterolaemia, the proportion of patients with a record of diabetes diagnosed after the age of 40 or lasting more than 10 years, or chronic kidney disease stages 3 to 5, was 19.4%, of whom 79.5% were prescribed statin therapy in accordance with current guidelines on primary prevention (Table 1). Likewise, the proportion of patients with hypercholesterolaemia and a history of cardiovascular disease, and therefore eligible for statin therapy in secondary prevention, was 10.5%. Among these patients, 81.4% were prescribed statins (Table 1).

### Association between statin prescription and stroke

The cause-specific hazard of stroke was assessed using a Cox proportional hazards model. To account for variation in the baseline hazard and to ensure that the proportional hazards assumption holds, the model was stratified by history of heart disease, and the follow-up was administratively censored at 79 years. The resulting dataset included 161527 patients with a record of hypercholesterolaemia (97.2%). The Schoenfeld test indicated no violation of the proportional hazards’ assumption at the 5% level for all variables fixed at baseline. Coefficients of the multivariable cause-specific hazard model are depicted in Figure 1. This model shows that statin prescription, female gender, and levels of deprivation from 3 to 5 (least deprived) were significantly associated with lower hazard of stroke. Black, Asian, and Mixed ethnicity (compared to White ethnicity), ever smoking status, hypertension and diabetes were significantly associated with higher hazard of stroke.

**Figure 1.**
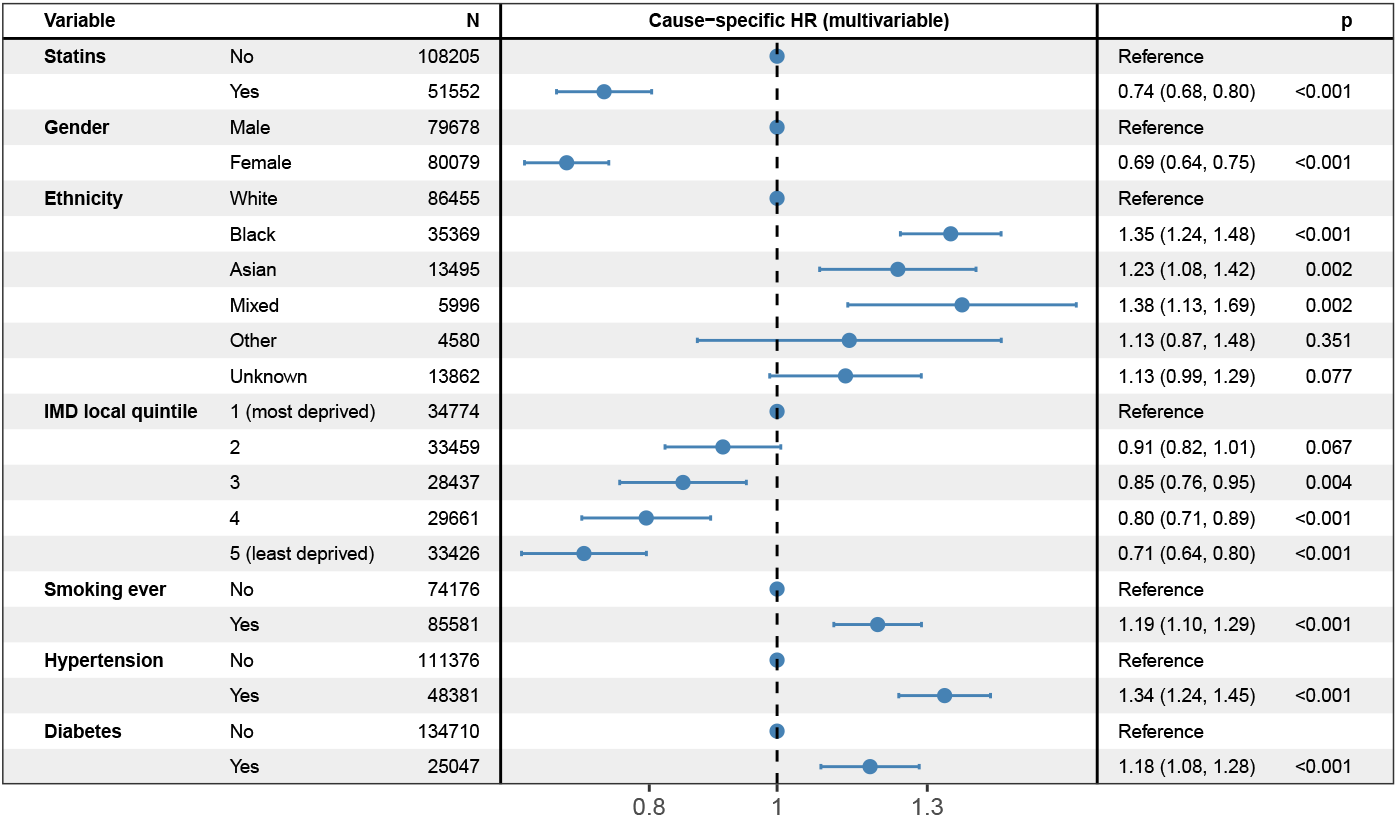
Multivariable cause-specific hazard ratio of statin prescription on stroke (allowing for time-varying exposure to statin prescription), adjusted for sociodemographics and stroke risk factors, and stratified by indicator of history of heart diseases, in patients with recorded hypercholesterolaemia, administratively censored at 79 years (n=159,757; number of events: 3,081; and 1,770 observations deleted due to missing covariates).

### Statin prescription in patients with hypercholesterolaemia

Figure 2 displays the multivariable odds ratios of statin prescription status associated with patients’ sociodemographic characteristics and stroke risk factors in patients with hypercholesterolaemia. As expected, statin prescription was strongly associated with stroke risk factors, particularly diabetes and hypertension, and, to a lesser extent, ever-smoking status. Black patients were 21% less likely to be prescribed statin therapy (odds ratio: 0.79; 95% CI: 0.76–0.82), while Asian patients were 65% more likely (odds ratio: 1.65; 95% CI: 1.57–1.74) compared to White patients. Finally, female patients were also 28% less likely to be prescribed statins than male patients (odds ratio: 0.72; 95% CI: 0.70–0.74). Additional file 1: Fig. S1 presents presents the calibration plot associated with the logistic regression displayed in figure This graph indicates overall good calibration, with predicted probabilities generally close to observed proportions across deciles. This visual impression corresponded to a Brier score of 0.13, with 0.22 representing the non-informative upper bound in our setting, and an area under the receiver operating characteristic curve of 87%, indicating good calibration and discrimination.

**Figure 2.**
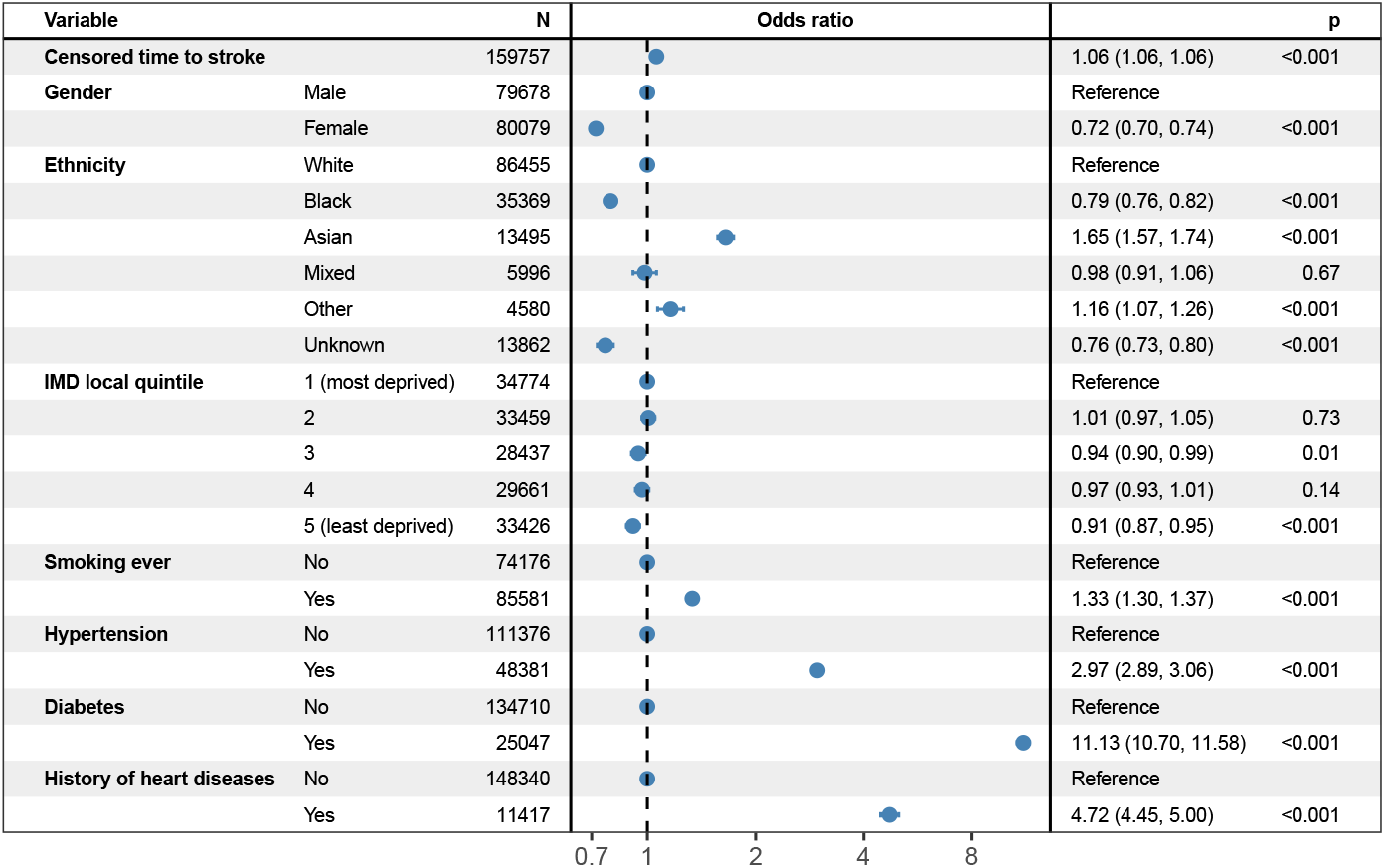
Multivariable odds ratio of statin prescription associated with sociodemographics and stroke risk factors in patients with hypercholesterolaemia (n= 164,319 and 1,805 observations deleted due to missing covariates)

## Discussion

In this study, we set out to better understand the association between the onset of stroke and statin prescription in patients with hypercholesterolaemia, using routinely collected electronic health records in a multiethnic, youthful, and urban population. Our results highlight the complexity of the relationship between hypercholesterolaemia and stroke, but also an association between statin prescription and reduced risk of stroke in patients with hypercholesterolaemia. Analysing the impact of statin prescription on stroke risk in observational studies is challenging. This is due mainly to indication bias where the likelihood of receiving statins, for instance, is itself associated with the underlying stroke risk factors [37]. To address this, we used a time-varying Cox proportional hazards model to estimate the cause-specific hazard of stroke according to statin prescription, adjusted for patients’ sociodemographic characteristics and stroke risk factors, while accounting for delayed records of statin prescription and potential competing events. In our analysis, patients were followed until the earliest of stroke, death, last recorded contact, or the point of administrative censoring. Consistent with our objective, this model allows estimation of the cause-specific hazard of stroke associated with statin prescription while appropriately handling death as a competing event for stroke [30–32], and avoids exposure misclassification and immortal time bias that occur when time-varying exposures are treated as fixed at baseline, whereby time before first prescription is incorrectly counted as exposed and artificially lowers the estimated hazard of stroke among treated patients [38]. This model showed a clear and significant downward effect associated with statin prescription on the hazard of stroke in patients with hypercholesterolaemia (cause-specific hazard: 0.74, 95% confidence interval: 0.68-0.80, p < 0.001, Figure 1).

These observations raise a number of questions, including the optimal prescription of statins in patients with hypercholesterolaemia and the role of statins’ pleiotropic effects [39] in delaying or preventing stroke onset in this population. In our study, 44.6% of patients with stroke had a history of hypercholesterolaemia, and among them, 61.1% were prescribed statins. This suggests that these patients were appropriately identified as being at higher risk for cardiovascular disease and accordingly were prescribed statin therapy, nearly twice the rate observed in other patients with hypercholesterolaemia (32.7%). While stroke could not be avoided in these patients, our data suggest that statin prescription is associated with a lower risk of stroke. Additionally, the post hoc analysis among patients with a history of stroke suggests that a substantial proportion could have benefited from statin therapy, potentially reducing their stroke risk. This perspective assumes, however, that statin therapy is further prescribed to patients with untreated hypercholesterolaemia, which represents 13.0% (n=110,522) of the registry population, but does not align with the current prevention policy. Another perspective reveals that only 1.5% of patients with untreated hypercholesterolaemia experienced a stroke, which is significantly lower than in patients with prescribed statins (Table 1), but is also slightly higher than the prevalence of stroke in the registry (1.2%). The multivariable analysis of patients’ sociodenmographic characteristics and stroke risk factors on statin prescription also shows that statins were prescribed according to current guidelines [22], that is, in patients with a history of cardiovascular disease and associated risk factors, including hypertension and diabetes. However, this analysis also reveals that female patients and patients of Black ethnicity were less likely to be prescribed statin therapy compared to male patients and patients of White and Asian ethnicity, respectively. This tendency may be due to implicit ethnicity-based biases by healthcare providers [40], misconceptions that women are at lower risk of cardiovascular disease [41, 42], or patient perceptions and beliefs regarding cardiovascular risk and statin therapy [43]. This tendency may also be linked to our previous observation that patients of Black ethnicity with hypercholesterolaemia had a higher risk of stroke compared to their counterparts of White and Asian ethnicity [44].

Of note, our analysis is based on recorded statin prescriptions. In other words, depending on treatment adherence and treatment gaps [26], prescriptions overestimate the actual exposure to statin therapy. As a consequence, it is likely that the true effect of statin therapy on stroke hazard is underestimated in our study. Conversely, depending on gender and ethnic patterns in statin therapy adherence, which appears to be lower in ethnic subgroups and women [45], it is also likely that the effect of gender and ethnicity on the risk of stroke is distorted. For instance, the higher risk of stroke observed in patients of non-White ethnicity is likely to be confounded with lower adherence in these patients, whereas the gender effect is likely to be underestimated as female patients exhibit both a lower risk of stroke and lower adherence to statin therapy. More generally, in addition to potential bias, including adherence bias, as discussed above, limitations and strengths of our study relate to both the use of routinely collected primary care electronic health records and the population under study. Although primary care electronic health records are widely regarded as a dependable data source and are increasingly utilised for research, surveillance, and public health planning [46, 47], the accuracy and completeness of recorded information may raise concerns. For example, records such as patients’ current smoking status, high-density lipoprotein (HDL) cholesterol, low-density lipoprotein (LDL) cholesterol, and QRISK3 score were not available in the electronic health records considered in this study.

Finally, the characteristics of the analysed population are important aspects to consider, as they may either support or limit specific study objectives. If the relative geographical and environmental uniformity within Lambeth borough, along with its diverse population, facilitates the identification of biases, such as inequalities in statin prescription practices, these characteristics should, however, be taken into account when attempting to generalise the study results to other settings.

## Conclusions

This study focused on statin prescription and the risk of stroke in patients with diagnosed hypercholesterolaemia. Overall, our results show that statin prescription is associated with a lower hazard of stroke in a real-life setting. Further insights reveal that guidelines for statin prescription are generally well followed by general practitioners, although post hoc analysis suggests that a small proportion of patients with untreated hypercholesterolaemia may have benefited from statin therapy. In line with NICE (updated) guidelines on statin prescription, our results further suggest that in uncertain or complex situations, and according to patients’ informed preference, comorbidity, sociodemographic characteristics, and ethnic background, statin therapy prescription may be considered even if patients do not present a 10-year cardiovascular risk higher than 10%. Finally, we recommend that further efforts be made to address disparities observed in statin prescription across gender groups and ethnic groups.

## List of abbreviations

HHD: History of Heart Disease
FH: Familial Hypercholesterolaemia
LDL-C: Low-Density Lipoprotein Cholesterol
NICE: National Institute for Health and Care Excellence
QRISK2: Cardiovascular Risk Assessment Tool version 2
QRISK3: Cardiovascular Risk Assessment Tool version 3
QOF: Quality and Outcomes Framework
IMD: Index of Multiple Deprivation
NIH: National Institutes of Health
HRA: Health Research Authority
CI: Confidence Interval
HDL: High-Density Lipoprotein
LDL: Low-Density Lipoprotein
NIHR: National Institute for Health and Care Research
AUC: Area Under the receiver operating characteristic Curve

## Declarations

### Consent for publication

Not applicable

### Data Availability

Data supporting this study can be obtained from the Lambeth Datanet subject to IRB approval, and are not publicly available.

### Authors’ contributions

MD designed the study (conceptualisation and methodology); AD acquired the data; MD carried out data curation; MD and AD conducted investigations; MD developed computer code and performed formal analysis, including data visualisation and validation; MD and AD interpreted the data. MD drafted the manuscript, and AD supervised the research. MD and AD reviewed and edited the final version of the manuscript. All authors read and approved the final manuscript.

### Ethics approval and consent to participate

Data were provided by the Lambeth DataNet upon approval for the analysis of fully anonymised data by the Lambeth Clinical Commissioning Group. All patients were informed through ‘Fair Processing Notices’ of the potential use of collected electronic health records in ‘secondary data analysis’ and were given the option to opt out of the data-sharing scheme. Accordingly, this study was exempt from ethics committee approval and individual consent requirements as per the National Institute for Health (NIH) Health Research Authority (HRA) guidelines for research using anonymised primary care data in the United Kingdom.

### Authors’ social media handles

BlueSky: @mdelord.bsky.social

X: @marc_delord

X: @lifecourse_KCL

### Additional files

Additional file 1: Fig. S1 and Table S1.

Fig. S1 – Calibration plot for the logistic regression modelling statin prescription as a function of covariates.

Table S1 – Patient characteristics according to stroke record. Additional file 2: STROBE Cohort Studies checklist.

## Acknowledgments

We would like to thank Dr Salma Ayis for her continuous help and support throughout the course of this work.

