## Supplementary material for "Statin Therapy and Stroke Risk in Patients with Hypercholesterolaemia: A population-based longitudinal study using electronic health records in South London"

### Supplementary materials

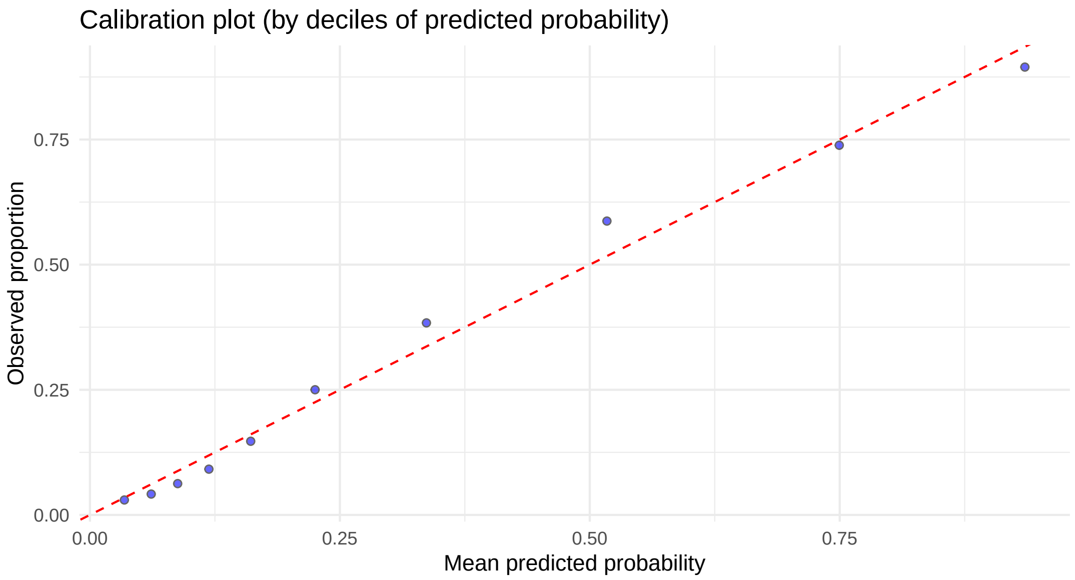


***Supplementary Figure S1:*** *Calibration plot: Calibration for the logistic regression where statin prescription is explain by age, gender, ethnicity, the local quintiles of the index of multiple deprivation, the ever smoking status, hypertension, diabetes, and history of heart diseases.*

Supplementary figure 1 depicts the calibration plot for the logistic regression displayed in figure 2 of the main manuscript. This graph is used to visually asses the agreement between predicted probabilities of statin prescription derived from the fitted model, and observed proportions of patients prescribed statin therapy, by decile of the predicted probabilities. A well-calibrated model should display points lying close to the 45-degree line, indicating good agreement between predicted and observed risks and showing that the logistic regression model fits the data adequately, yielding reliable predicted probabilities.

### ***Supplementary Table S1:*** *Patient characteristics according to stroke record*

| | | Stroke | | |
|---|---|---|---|---|
| Patient characteristics | | Not recorded | Recorded | Total |
| Total N (%) | | 161734 (97.4) | 4390 (2.6) | 166124 |
| Age at end of follow-up | Median (IQR) | 56.1 (22.0) | 71.3 (20.5) | 56.2 (22.2) |
| Gender | Male | 79923 (49.4) | 2194 (50.0) | 82117 (49.4) |
| | Female | 81808 (50.6) | 2196 (50.0) | 84004 (50.6) |
| Ethnicity | White | 88094 (54.5) | 2299 (52.4) | 90393 (54.4) |
| | Black | 35208 (21.8) | 1167 (26.6) | 36375 (21.9) |
| | Asian | 13545 (8.4) | 326 (7.4) | 13871 (8.3) |
| | Mixed | 5997 (3.7) | 133 (3.0) | 6130 (3.7) |
| | Other | 4597 (2.8) | 66 (1.5) | 4663 (2.8) |
| | Unknown | 14293 (8.8) | 399 (9.1) | 14692 (8.8) |
| IMD local quintile | 1 (most deprived) | 34680 (21.7) | 1137 (26.1) | 35817 (21.8) |
| | 2 | 33548 (21.0) | 948 (21.7) | 34496 (21.0) |
| | 3 | 28428 (17.8) | 701 (16.1) | 29129 (17.7) |
| | 4 | 29744 (18.6) | 683 (15.7) | 30427 (18.5) |
| | 5 (least deprived) | 33561 (21.0) | 892 (20.5) | 34453 (21.0) |
| Age at record of Hypercholesterolaemia | Median (IQR) | 46.8 (18.1) | 63.1 (19.4) | 47.2 (18.4) |
| Statins | Not prescribed | 108813 (67.3) | 1709 (38.9) | 110522 (66.5) |
| | Prescribed | 52921 (32.7) | 2681 (61.1) | 55602 (33.5) |
| Hypertension | No | 112064 (69.3) | 1427 (32.5) | 113491 (68.3) |
| | Yes | 49670 (30.7) | 2963 (67.5) | 52633 (31.7) |
| Diabetes | No | 136117 (84.2) | 3087 (70.3) | 139204 (83.8) |
| | Yes | 25617 (15.8) | 1303 (29.7) | 26920 (16.2) |
| CKD 3-5[a] | No | 150932 (93.3) | 3665 (83.5) | 154597 (93.1) |
| | Yes | 10802 (6.7) | 725 (16.5) | 11527 (6.9) |
| Smoking ever | No | 74869 (46.3) | 1795 (40.9) | 76664 (46.1) |
| | Yes | 86865 (53.7) | 2595 (59.1) | 89460 (53.9) |
| Obesity (moderate to severe) | No | 104936 (64.9) | 2749 (62.6) | 107685 (64.8) |
| | Yes | 56798 (35.1) | 1641 (37.4) | 58439 (35.2) |
| History of heart disease[b] | No | 148282 (91.7) | 3109 (70.8) | 151391 (91.1) |
| | Yes | 13452 (8.3) | 1281 (29.2) | 14733 (8.9) |
| History of cardiovascular disease[d] | No | 146113 (90.3) | 2536 (57.8) | 148649 (89.5) |
| | Yes | 15621 (9.7) | 1854 (42.2) | 17475 (10.5) |
| Diabetes*[c] or CKD 3-5 | No | 131263 (81.2) | 2692 (61.3) | 133955 (80.6) |
| | Yes | 30471 (18.8) | 1698 (38.7) | 32169 (19.4) |

[a] Chronic Kidney Disease Stage 3-5

[b] Myocardial infarction, Heart Failure, Coronary Heart Disease, Atrial Fibrillation

[c] History of Heart Disease, Transient Ischemic Attack, Peripheral Arterial/Vascular Disease

[d] Diabetes after 40 years old or for more than 10 years
